# Selphi: Empowering GWAS Discovery through Enhanced Genotype Imputation

**DOI:** 10.1101/2023.12.18.23300143

**Authors:** Adriano De Marino, Abdallah Amr Mahmoud, Sandra Bohn, Jon Lerga-Jaso, Biljana Novković, Charlie Manson, Salvatore Loguercio, Andrew Terpolovsky, Mykyta Matushyn, Ali Torkamani, Puya G. Yazdi

## Abstract

**Motivation:** Genotype imputation is a powerful tool for inferring missing genotype data in large-scale genomic studies. Over the last two decades, multiple research groups have developed a number of imputation algorithms, which continue improving in speed and overall accuracy. However, accurate imputation of rare and infrequent variants remains a challenge.

**Results:** Here we present Selphi, a novel genotype imputation algorithm based on the Positional Burrow Wheeler Transform (PBWT) and a new heuristic method for haplotype selection based on identity by descent (IBD). When compared to state-of-the-art methods Beagle5.4, IMPUTE5, and Minimac4, Selphi showed a higher accuracy in 1000 Genome Project and TOPmed datasets, across all super-populations and allele frequencies. Similarly, Selphi performed better than Beagle5.4 in the UK Biobank dataset, which translated into improved GWAS discovery and more accurate polygenic risk scores. Selphi’s improvements in imputation accuracy, especially for rare and low frequency variants, promises to boost the power and accuracy of downstream genomic applications.

**Availability and implementation:** Selphi code is available at GitHub: https://github.com/selfdecode/rd-imputation-selphi. Additionally, we offer an applet that allows convenient testing of the Selphi code on the UKB RAP platform.

## Introduction

Genomic medicine has grown rapidly since the completion of the Human Genome Project (HGP) two decades ago, promising to deepen our knowledge of disease pathology, improve diagnostic speed and accuracy, and enable targeted disease treatment and preventive therapy (Mega et al. 2015, Natarajan et al. 2017, Damask et al. 2020, Marston et al. 2020, Pereira et al. 2021, Klarin and Natarajan 2022, Mishra et al. 2022). However, as whole genome sequencing (WGS) is still prohibitively expensive, especially when it comes to large-scale population-wide screening, a lot of academic and direct-to-consumer efforts rely on array-based SNP genotyping and low-coverage WGS (lcWGS). These approaches are cost-effective, but their accuracy is limited by the accuracy of imputation methods used to fill in the gaps in these datasets. Therefore, imputation has a profound impact on all downstream applications when using genotyping and lcWGS datasets, such as detecting associated variants in genome-wide association studies (GWAS) or calculating polygenic risk scores (PRS) (Pasaniuc et al. 2012, Homburger et al. 2019, Chen et al. 2020, Nguyen et al. 2022, Appadurai et al. 2023).

Over the last twenty years, multiple groups have developed and published different imputation methods, the majority of which are based on the Li and Stephens Hidden Markov Model (HMM) (Scheet and Stephens 2006, Fuchsberger et al. 2015, Das et al. 2016, Browning et al. 2018, Delaneau et al. 2019, Rubinacci et al. 2020). However, they still suffer in accuracy when imputing rare variants (Herzig et al. 2018, Sariya et al. 2019, De Marino et al. 2022) This is of note because rare variants can be highly informative and of great medical significance (Bomba et al. 2017, Momozawa and Mizukami 2021, Weiner et al. 2023). Recently, efforts have been made to address this by broadening the reference panel either by creating cohort-specific imputation reference panels (Mitt et al. 2017, Sun et al. 2022, Xu et al. 2022), or adding whole exome sequencing data into the reference panel (Wuttke et al. 2023). However, improving rare variant imputation by tackling the algorithm itself has not yet been convincingly demonstrated. Here, we describe a new imputation tool that does just that by identifying and giving priority to potential identity by descent (IBD) segments.

Selphi is a tool that improves imputation by using a heuristic algorithm to select potential IBD haplotype segments coupled with a Positional Burrow Wheeler Transform (PBWT) (Durbin 2014). We compare Selphi to state-of-the-art imputation methods Beagle5.4 (Browning et al. 2018), IMPUTE5 (Rubinacci et al. 2020), and Minimac4 (Das et al. 2016) using the 1000 Genomes Project (1000 Genomes Project Consortium 2015) and TOPmed (Taliun et al. 2021) datasets. Next we impute UK biobank (Bycroft et al. 2018) genotyping data using Selphi and the next most accurate model, Beagle5.4 to demonstrate the applicability of Selphi to large-scale datasets. Finally we use this imputed data to improve downstream GWAS and PRS.

## Methods

### Selphi

#### Model Overview

Selphi was developed in Python and C and uses vectorized matrix functionalities of numpy for efficient processing in large-scale data environments. It operates under the assumption that both reference and target genotypes are phased and non-missing. The imputation process relies on the concept of identity by descent (IBD), identifying chromosome segments inherited from a common ancestor uninterrupted by recombination events, which allows for the transfer of un-genotyped alleles from reference to target haplotypes (Figure 1).

**Figure 1.**
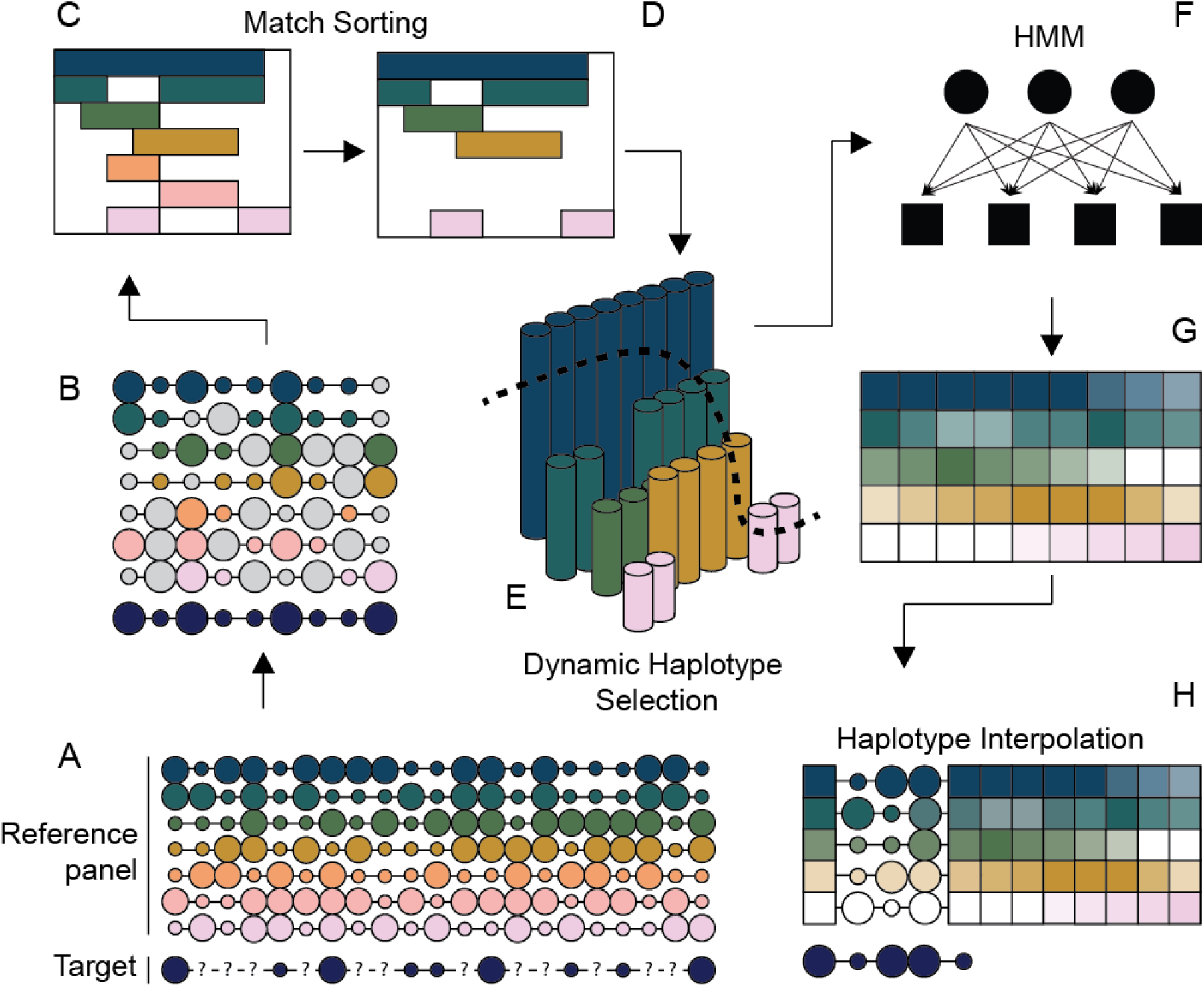
Selphi workflow. (A) The first step in the workflow involves merging the reference panel and target data into a unified PBWT data structure. (B) Subsequently, the algorithm scans the reference panel searching for matches to reference haplotypes of a minimum length L. (C–D) At each marker, the algorithm retains the longest matches, prioritizing haplotypes with more total matches across the chromosome. (E) A dynamic haplotype selection step follows, where the matches are mapped and filtered to adjust the number of retained matches at each marker, based on the distribution of match lengths. (F) The HMM forward-backward algorithm is employed. (G) Transitions between variant states are utilized to compute weights for each haplotype at each marker. These weights aid in determining the significance of each haplotype within the population. (H) The final step involves the interpolation of allele probabilities with the haplotypes from the reference panel.

#### PBWT

For improved precision, Selphi integrates the Positional Burrow Wheeler Transformation (PBWT) (Durbin 2014), adept at identifying the longest haplotype matches between the target and reference sequences. PBWT’s primary objective is to arrange haplotypes in a reversed prefix order, a mechanism that markedly simplifies the identification and matching of haplotypes across datasets. The model also includes a heuristic Identity by Descent (IBD) selection at each genotyped marker, crucial for filtering out coincidental Identity by State (IBS) matches.

The PBWT algorithm initiates with the construction of a positional prefix array. This array is essentially a sequence of haplotype indices, arranged such that the haplotypes are sorted in reverse prefix order at a given position, denoted as *n*. To achieve this, two distinct vectors, of length *M,* are created for each genotype marker at position *n*. One vector is responsible for holding the indices of haplotypes, sorted according to their reversed prefix order. The other vector tracks the index where the last match for each haplotype began, essentially marking the starting point of each haplotype match. This helps facilitate quick and memory-efficient pairwise comparisons between all haplotypes in the reference panel.

In the context of Selphi’s imputation process, the reference panel denoted as *X* with X {0,1}MN and the target haplotype *T* with T {0,1}1N are defined within a certain genomic structure. Here, *N* represents the total number of genotyped variants. The reference panel X and the target T are aligned such that they share a common set of markers, with the reference panel not necessarily containing a complete marker set but only those that overlap with the target.

The first crucial step in Selphi’s imputation process involves the computation of forward matches. This is achieved by accumulating previous matches for the same haplotype until a mismatch occurs, at which point the match count resets to zero. This mechanism is integral to the PBWT and is detailed in equation (1), describing the creation of BI data structures.

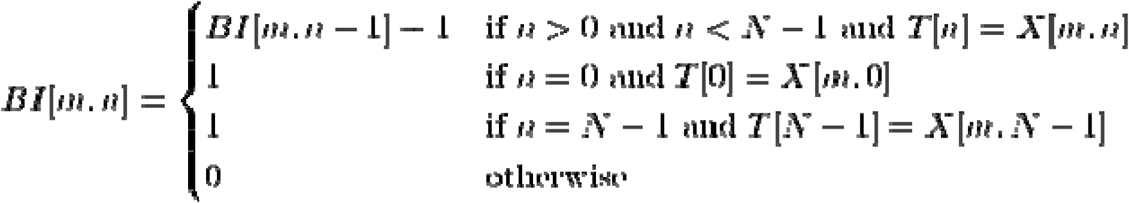

**Equation 1**: Calculation of forward matches (BI data structures). This mechanism involves accumulating previous matches for the same haplotype until a mismatch occurs, at which point the match count resets to zero. This process is integral to PBWT.

The algorithm then proceeds to compare each target haplotype against every reference haplotype at each variant. A match is recorded when there is a divergence, provided that the total length of the match exceeds a pre-set threshold, typically a minimum of five consecutive variants. This threshold ensures that only significant matches are considered, enhancing the accuracy of imputation. The matches are then organized into a sparse matrix format, which is particularly suited for handling data with a high proportion of zero values, common in genomic matrices. The sparse matrix, encapsulating the essential match data, is then saved as a .npz file for downstream use.

#### Haplotype selection

The haplotype selection process begins with the construction of a custom match matrix, which serves as a structured representation of consecutive haplotype matches identified through the PBWT. Each entry in this matrix represents the length of consecutive matches between a target haplotype and reference haplotypes. To refine this selection, Selphi constructs a filtering mask—a secondary matrix that delineates the maximum length of matches at each genomic marker. In this matrix, for each marker, the k haplotypes with the longest matches are retained, effectively filtering out less likely haplotypes and thus narrowing down the potential candidates for imputation.

With this filtered matrix, Selphi then assigns weights to each match, incorporating both (i) the length of individual matches and (ii) the aggregated matching performance across all markers. These weighted values are stored in a weighted matrix (*WH*) and used in determining the haplotype’s contribution. Each haplotype is then weighted according to equation (2).

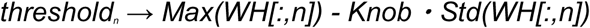

**Equation 2**: Computation of the haplotype match weighting threshold (*threshold_n_*). The formula involves subtracting the product of the knob parameter and the standard deviation (*Std*) of the weighted matrix (*WH*) at a specific genomic marker from the maximum value at that marker (*Max(WH[:,n])*).

Equation 2 computes a threshold (*threshold_n_*) for each genomic marker *n*, where:

- *Max(WH[:,n])* is the maximum weighted match length at marker *n*,
- Knob is an adjustable scaling parameter proportionate to the distance between the mean weighted match length at marker n and the maximum match length,
- *Std(WH[:,n])* is the standard deviation of the weighted match lengths at marker *n*.

The threshold dynamically adjusts based on the distribution of match lengths at each marker. Markers with longer matches have a higher threshold, and haplotypes with match lengths that fall below this threshold are excluded from the imputation process. This mechanism allows Selphi to focus on high-confidence matches, improving the overall accuracy of the imputation. When high-confidence matches are not present, Selphi includes a broader sampling of haplotypes in the imputation process.

Haplotype selection is conducted across the entire chromosome without segmenting it into windows. This is a distinguishing feature of our method, contrasting with others that divide the genome into smaller windows (Browning et al. 2018, Rubinacci et al. 2020). This comprehensive approach ensures the conservation of essential data derived from the pairing of target sequences with the reference panel. Moreover, it avoids the imputation inaccuracy near window boundaries, a known limitation in methods employing short, non-overlapping windows.

Our technique circumvents the potential loss of continuity and the need for overlapping windows, thus enhancing the integrity and consistency of the imputation results.

#### Dynamic Thresholding and the Knob Parameter

A critical element of the haplotype selection process is the knob parameter, which is an adjustable factor used to control the stringency of haplotype selection. The knob parameter is calibrated against the average match length at each marker, normalized between 0.2 and 3. This normalization allows for fine-tuning the sensitivity of the selection process based on the data distribution:

- Lower knob values are applied when the number of potential matches is high. This stricter criterion is necessary to filter out weaker haplotype matches and ensure that only the strongest candidates, those likely to represent true Identity by Descent (IBD), are retained.
- Higher knob values are used when there are fewer matches. This relaxed threshold prevents the exclusion of valid haplotypes in cases where genuine matches may be shorter due to data sparsity or variability in the genetic sequences.

By dynamically adjusting the threshold for haplotype inclusion, the knob parameter mitigates the influence of Identity by State (IBS) matches—instances where haplotypes share alleles by chance rather than through a common ancestor. This tunable selection process ensures that the imputation algorithm remains both sensitive and specific, maintaining a balance between false positives and false negatives.

#### Imputation

The imputation component of Selphi utilizes a modified version of the Li Stephens Hidden Markov Model (HMM) (Li and Stephens 2003, Li et al. 2009). In Selphi’s adaptation of HMM, the *hidden states*, which are not directly observable, are represented by haplotypes at specific loci across the genome, denoted by pairs of indices (*m,n*), with *m* indexing the haplotype within the reference panel and *n* designating the particular genetic marker in question.

Our method diverges from the standard use of the forward-backward algorithm for imputation, primarily because of how we define transition probabilities. We permit a complete transition probability for a move from one hidden state to a subsequent state, provided that the haplotype’s position in the reference panel remains consistent (this condition is depicted as *H_m_ + 1 = H_m_*). *H_m_* stands for a hidden state at marker index m. Here, *hidden state* refers to a specific haplotype in the reference panel of haplotypes, and m is the index of that haplotype in the reference panel. In this condition we allow the model to have a full transition probability (equal to 1) from one state to another state when the haplotype does not change from one marker to the next.

The exact probability of such transitions is outlined in equation (3), where *Ne* denotes the effective population size, which is typically assumed to be 1,000,000. dm signifies the recombination distance, which we derive through linear interpolation of the distances provided by a publicly available genetic map (genetic map). Finally, *NumHid* corresponds to the total number of hidden states.

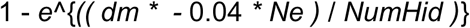

**Equation 3**: Transition probability between states using recombination rate.

The forward-backward algorithm is used to estimate the probabilities of missing genetic marker data. This process has been optimized by implementing the forward-backward algorithm using sparse matrices for both forward and backward passes, which considerably reduces computational load. The transition probabilities are then used to infer the most likely haplotypes given the observed genotypes.

For computational efficiency, Selphi processes each haplotype in parallel, dedicating a computing core to each target haplotype. This parallel processing extends to the interpolation of reference states, following a method akin to that used in Beagle5.4 (Browning et al. 2018) and IMPUTE5 (Rubinacci et al. 2020), where linear interpolation between two boundary probabilities is employed to compute the reference states. The cumulative probabilities for both the reference and alternate alleles are then computed at each marker, culminating in the imputed genetic profile.

#### Sparse reference format (.srp)

Efficient interpolation requires rapid retrieval of selected haplotypes at markers within the interpolation window. Selphi includes a customized tool for compressing large reference panels into chunked sparse matrices, enabling rapid access of reference panel data with a smaller storage footprint than a compressed VCF (Danecek et al. 2011). Reference panel haplotypes are converted to sparse matrices, each containing a preset number of markers. The sparse matrices are compressed with Zstandard compression and organized within a zip archive, allowing rapid loading into memory. Once loaded, sparse matrices are cached in memory until they are no longer accessed, eliminating disk latency as Selphi moves down the chromosome. The chunked storage format also allows Selphi to parallelize imputation across the chromosome without loss of performance.

Selphi offers good flexibility by allowing generation of the srp format from both compressed/uncompressed VCF/BCF and XSI reference formats (Wertenbroek et al. 2022), enhancing its adaptability to diverse data sources. This versatility empowers Selphi to seamlessly handle various reference data types. The .srp format’s inherent flexibility streamlines the process, ensuring smooth and reliable imputation even in scenarios with a large number of samples and when the data size of a compressed VCF reference panel could be problematic to handle.

### Datasets

#### 1000 Genomes Project

The 1000 Genomes Project 30x dataset contains phased sequences of 3,202 individuals sampled from 26 different populations. We selected all the individuals without relatives in the dataset to test imputation in unrelated individuals. The filtering was executed using the pedigree file available at (http://ftp.1000genomes.ebi.ac.uk/vol1/ftp/technical/working/20130606_sample_info/20130606_g1k.ped). Individuals that had a Family ID that diverged from the Individual ID were selected and used as our reference panel while individuals with the same Family ID and Individual ID were used as the target dataset for imputation. This filtering ensures that there are no related individuals between the target and reference panel that could inflate imputation results. The number of samples in the reference panel was 2401. The final number of target samples was 801, belonging to 12 out of the 26 populations found in the dataset (Figure S4). All analysis used the hc-WGS 30x version of the 1000 Genomes Project (1KG) (1000 Genomes Project Consortium 2015).

We used the following filtering criteria for all variants: (i) only variants with FILTER=PASS were retained; (ii) variants with genotype missingness below 5% were included; (iii) variants passing the Hardy-Weinberg equilibrium (HWE) test, indicated by an HWE *P* value greater than 10^-10^ in at least one of the five super-populations, were kept; (iv) variants with a Mendelian error rate of 5% or lower were considered; and finally, (v) variants with a minor allele count (MAC) of 2 or higher were included.

To assess imputation accuracy we masked a portion of the markers to simulate genotyping data. We limited the 1000 Genomes reference data to markers that had at least one minor allele copy in the reference panel, and we masked markers not found on the Illumina GSA chip array (Table S4). This masking process was applied to all chromosomes using the GSA chip array as reference (GSA v3 by Illumina).

#### TOPMED Dataset

To compare imputation performance against a more diverse reference panel, we assembled a larger, ethnically and ancestrally diverse reference panel using WGS data from 32 studies (48 consensus groups) available through the NHLBI TOPMed (Trans-Omics for Precision Medicine) Program (Taliun et al. 2021), encompassing 90,897 participants. We considered the Freeze 8, GRCh38 version of TOPmed data, which was the latest version with all consent groups bearing the same number of variants. For details regarding the processing of TOPmed Freeze 8, see (https://topmed.nhlbi.nih.gov/topmed-whole-genome-sequencing-methods-freeze-8). TOPmed data were made available as gVCF files through the database of Genotypes and Phenotypes (dbGaP). All study names and IDs are listed in Table S5. We focused on chromosome 20 and performed haplotype phasing using Beagle5.4 (Browning et al. 2018), which is particularly optimized for large datasets like TOPMed.

From the TOPMed dataset, we selected a subset of 5,000 samples, coming from the Multi-Ethnic Study of Atherosclerosis (MESA) (Bild et al. 2002, Olson et al. 2016) for our imputation experiments (White-Caucasian 1634, Black-African-American 930, Hispanic 862, Chinese-American 535). These selected samples were unrelated to each other within the dataset. The remaining 85,897 WGS samples from the TOPMed dataset served as a comprehensive reference panel.

The quality control (QC) steps were executed as follows: Initially, we split multi-allelic variants into bi-allelic forms using BCFtools (Danecek et al. 2021). The subsequent filtering of SNPs and indels was based on several criteria: (i) a Hardy-Weinberg equilibrium *P* value less than 10^-30^, (ii) more than 5% missing data among individuals (based on a GQ score = 0), (iii) abnormal heterozygosity rates, defined as less than 0.5 or greater than 1.5, (iv) alternative alleles with an AA-score below 0.5, (v) variants where the FILTER field was not ‘PASS’, (vi) kept only biallelic SNPs. These QC measures are crucial for ensuring the reliability of subsequent analyses and were automated within the TOPMed data processing framework.

A total of 46 phased VCF files – one for each consensus group, excluding two from MESA - were then merged. The final reference panel for chr20 thus assembled consisted of 85,897 samples and 17,900,635 biallelic SNPs. The reference was also converted into formats appropriate for each imputation tool (.bref3 format for Beagle5, .m3vcf for Minimac4 and imp5 for IMPUTE5). For imputation validation, we used genotype data derived from a masking of MESA samples, using the GSA SNP array.

#### UK Biobank

We used the 150,119 WGS data jointly called with GraphTyper v2.7.1 (Eggertsson et al. 2017), available as pVCF files on the UK Biobank RAP (Rubinacci et al. 2023). We selected all autosomal chromosomes and conducted haplotype phasing using Shapeit v4.2.2 (Delaneau et al. 2019). The quality control process was carried out as follows: initially, multi-allelic variants were decomposed into bi-allelic variants using BCFtools (Danecek et al. 2021). Subsequently, SNPs and indels were filtered based on several criteria: (i) a Hardy-Weinberg p-value lower than 10^-30^, (ii) over 5% of individuals with missing data (GQ score = 0), (iii) an excess of heterozygosity, measured as less than 0.5 or greater than 1.5, (iv) alternative alleles with an AA-score below 0.5, and (v) variant sites where the FILTER tag did not match PASS. We selected a subset of 50,000 samples with White British ancestry from the UK Biobank dataset. These samples were unrelated to any other individual in the dataset and had Axiom SNP array data available for imputation experiments, making them the target samples. The remaining 100,119 Whole-Genome Sequencing (WGS) samples from the UK Biobank were utilized as the reference panel. Phased Axiom genotype data have been downloaded from the UK Biobank study conducted by Bycroft et al. (2018) (Bycroft et al. 2018). Subsequently, the data was lifted over to the GRCh38 human reference genome, with strand flips discarded, resulting in a dataset comprising 657,354 autosomal markers for 487,442 samples (Table S6). After liftover, approximately 99.8% of the original variants were retained for further analysis.

#### Benchmarking

We analyzed the autosomal chromosomes from the 1KG reference panel to explore the distribution of the selected states in our imputation experiments. The 801 unrelated target samples were imputed against the remaining haplotypes in the reference panels. We compared the accuracy of Selphi with the most up-to-date versions of Beagle5.4 (Browning et al. 2018), IMPUTE5 (Rubinacci et al. 2020) and Minimac4 (Das et al. 2016), using default parameters for each program. We used the true genetic map for analyses for Beagle5.4, IMPUTE5 and Selphi for real data imputation. Minimac4 does not require a genetic map, as recombination parameters are estimated and stored when producing the m3vcf format input file for the reference data.

The accuracy of the methods was assessed by comparing the imputed allele probabilities to the true (masked) alleles, as previously described (De Marino et al. 2022). Markers were binned into bins according to the minor allele frequency of the marker in the reference panel. For each bin we also calculated the squared correlation (*r*^2^) between the vector of all the true (masked) alleles and the vector of all posterior imputed allele probabilities, the number of errors in concordance with the true masked allele, and the imputation quality score (IQS)Precision and Recall as the F-score (Browning and Browning 2009, Lin et al. 2010). For the imputation accuracy evaluation, we have rebuilt a faster version of the tool Simpy to obtain all evaluation metrics (De Marino et al. 2022). All imputation analyses for the 1000 Genomes Project (1KG) were conducted on an AWS EC2 instance featuring a 107-vCPU computer equipped with Intel Xeon Platinum 8171M CPU processors and 753 GB of memory.

For the TOPMed dataset, we focused our analysis solely on chromosome 20 for efficacy, following the same exact methodology stated previously. All computations for TOPMed were performed at Scripps HPC (High Performance Computing) facility through a Singularity image (Kurtzer et al. 2017), using a variable number of 16-CPU nodes equipped with 128Gb RAM.

For the UK Biobank (UKB) dataset, a similar approach was employed. The imputation experiments were conducted on the UKB RAP platform. To execute Selphi on the UKB RAP platform, our software was developed as a single applet on the dnanexus platform. These applets were run with distinct hardware configurations, employing virtual machines (VMs) tailored to meet the minimum hardware requirements specific to each chromosome being imputed.

#### GWAS analysis

Following the imputation of all autosomal chromosomes for the entire cohort of 50,000 individuals of white British ancestry from the UK Biobank, we selected 50 phenotypes (Table S7) with less than 10% missing data across anthropomorphic traits and blood measurements in our call set for further analysis. To assess associations between the selected phenotypes and the imputed call sets, we utilized plink2 (Chang et al. 2015) with default parameters, incorporating sex, age, and the first 10 principal components (PCs) as covariates. We analyzed the hc-WGS dataset, along with two datasets imputed by Beagle5.4 and Selphi. Our locus selection criteria involved two key factors: (i) we focused on genome-wide significant loci with *P* values less than 5e-08 reported by the NHGRI Catalog of published GWAS (release 2023-08-26) (MacArthur et al. 2017), and (ii) we considered the strongest signal per locus (±100□kb genomic region) to select independent loci. For the analysis, we exclusively considered imputed variants, removing those present in the axiom array. To compare beta values (slope) and *P* values (significance) between the imputed set and the results obtained with the hc-WGS set, we adopted two approaches: (1) using absolute beta values and (2) employing the negative logarithm of the *P* value on a logarithmic scale to address low and highly significant *P* values (for at least nominally significant associations, *P* < 0.05). In evaluating concordance (*r*^2^) in the correlation of imputed vs. hc-WGS association values, we assessed how well the data fit the 1:1 identity line.

#### PRS analysis

In the final phase of our study, we utilized seven GWAS phenotypes mentioned earlier to generate PRS scores, including atrial fibrillation, asthma, hypertension, type 2 diabetes, height, apolipoprotein B levels, and calcium. To facilitate the analysis, quantitative traits were transformed into binary categories: short stature was defined as the lowest 10% of individuals based on height, accounting for sex; hypercalcemia was characterized by calcium levels exceeding 2.6 mmol/L; and high ApoB was designated for levels surpassing 1.3 g/L. We generated summary statistics by meta-analyzing existing external datasets collected by the GWAS Catalog (Table S8). We implemented clumping plus thresholding models, exploring various parameter values as detailed by Privé et al. (Privé et al. 2019). Specifically, we investigated squared correlation thresholds of clumping within {0.01, 0.05, 0.1, 0.2, 0.5, 0.8, 0.95}, base sizes of clumping windows within {50, 100, 200, 500} divided by *r*^2^ of clumping (parameter 1), a sequence of 50 thresholds on *P* values between the least and most significant values on a log-log scale, and 13 minor allele frequency (MAF) threshold filters ranging from 0.001 to 0.1. During the training phase, we assessed a total of 18,200 PRS models per phenotype and selected the most accurate one for each callset as the optimal hyperparameters. The 50,000 individuals from the UK Biobank (UKBB), utilized in the GWAS power analysis, were divided into 30,000 for training and 20,000 for testing the PRS models. The assessment of PRS accuracy involved two key measures: (i) relative risk, defined as the ratio of the percentage of cases found between the fifth quintile (individuals with high PRS) and the first quintile (individuals with low PRS) of the PRS distribution; and (ii) area under the curve (AUC).

## Results

### Selphi Improves Rare Variant Imputation

To benchmark Selphi’s performance, we first compared its accuracy against that of Beagle5.4, IMPUTE5 and Minimac4, using chromosomes 1-22 of the 1000 Genomes Project dataset (Figure 2a) (1000 Genomes Project Consortium 2015). This dataset has been widely used as a gold-standard dataset for testing imputation accuracy (Browning et al. 2018, Sariya et al. 2019, De Marino et al. 2022,). Selphi had an overall concordance of 0.9941 (Table S1), with an improvement in accuracy of 4.2% to 27.3% per sample. Our model achieved the best results with the lowest number of errors across all minor allele frequency intervals and all ancestral backgrounds (Figure 2a; Table S2). It performed exceedingly well for rare (MAF 0.05-2%) and particularly for ultra-rare (MAF 0.05-0.1%) variants, with an average improvement of 13% and 21%, respectively. The improvement was pronounced in the East Asian and African super- populations, with 16.4% and 4% improvement, respectively, compared to the next best model Beagle5.4. We additionally assessed the accuracy of each method using the following metrics: squared correlation (*r*2), concordance (P0) and imputation quality score (IQS) (Figure S1; Tables S1-2) (Browning and Browning 2009, Lin et al. 2010). Selphi remained the best method across all evaluated metrics across all ancestries (Figure S1, Table S1, Table S2).

**Figure 2.**
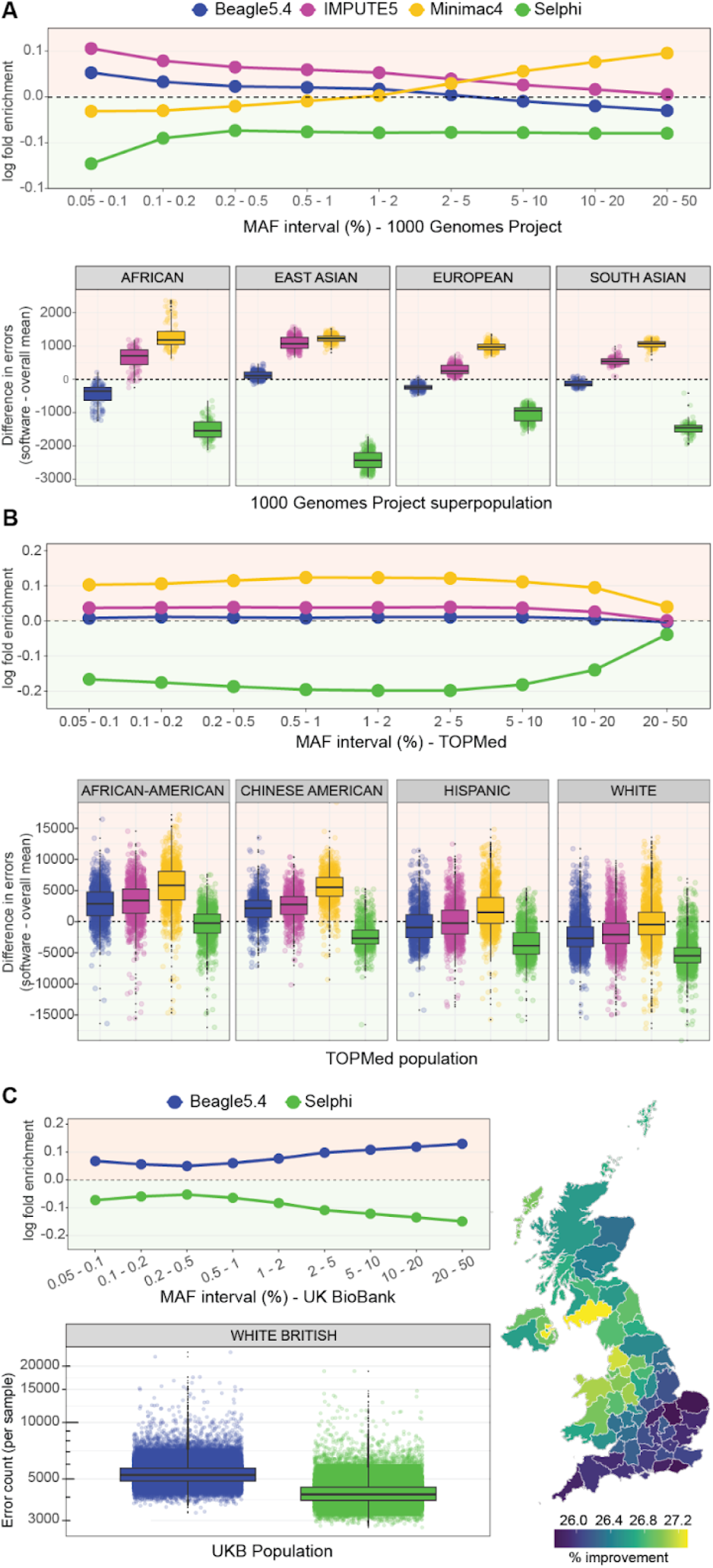
Selphi improves rare and low frequency variant imputation. (A) Relative enrichment (red background) and depletion (green background) of error counts with respect to average for Beagle5.4 (blue), IMPUTE5 (magenta), Minimac4 (yellow) and Selphi (green) binned by minor allele frequencies (MAFs) and by different populations across chromosomes 1-22 of the 1000 Genomes Project (1KG) 30x reference panel. (B) Relative enrichment (red background) and depletion (green background) of error counts with respect to average for Beagle5.4 (blue), IMPUTE5 (magenta), Minimac4 (yellow) and Selphi (green) binned by minor allele frequencies (MAFs) and by different populations for chromosome 20 of the TOPmed dataset. (C) Relative enrichment and depletion of error counts with respect to average and error count per sample for chromosomes 1-22 of the UK Biobank dataset. Map shows improvement in imputation accuracy across UK counties against Beagle5.4.

Next, we benchmarked Selphi’s accuracy using chromosome 20 of TOPmed (Taliun et al. 2021), a large, ethnically and ancestrally diverse dataset that is increasingly used to improve imputation accuracy, especially in admixed populations (Huerta-Chagoya et al. 2023). 5,000 samples from the TOPMed dataset’s Multi-Ethnic Study of Atherosclerosis (MESA) were imputed against the remaining 85,897 high coverage WGS (hc-WGS) TOPMed samples as the reference panel. Selphi again achieved the best results with the lowest number of errors, with an average improvement of 27.1% for rare variants (MAF 0.05-2%) (Figure 2b; Figure S2, Table S1, Table S3).

Finally, to demonstrate its applicability to larger datasets, we benchmarked Selphi against Beagle5.4, the next most accurate model in our analysis, using the UK Biobank dataset (Bycroft et al. 2018). We imputed chromosomes 1-22 of 50,000 samples classified as White British. Selphi performed better than Beagle5.4 for all MAFs (Figure 2c). On average, Selphi accomplished a ∼25% increase in concordance over Beagle5.4, with an improvement of 13.4% for rare variants (MAF 0.05-2%). Notably, for the MAF interval of 20-50%, Selphi made around 20,000 fewer errors per sample, achieving 33.4% improvement (Table S1, Table S4).

### Selphi Improves GWAS Variant Discovery and PRS

To ascertain that improved imputation accuracy would boost GWAS variant discovery, we used 50,000 unrelated White British samples from the UK Biobank that possessed both genotyping and hc-WGS information, and imputed their genotyping data using Selphi and Beagle5.4. Next, we conducted GWAS for 50 distinct traits using the imputed datasets. Selphi yielded results in closer alignment with the hc-WGS data, especially for rare variants (Figure 3a-c, Figure S3). Finally, we used the GWAS results to create polygenic risk scores (PRS) for seven different phenotypes. Imputation by Selphi produced PRSs in closer alignment to that of hc-WGS (Figure 3d).

**Figure 3.**
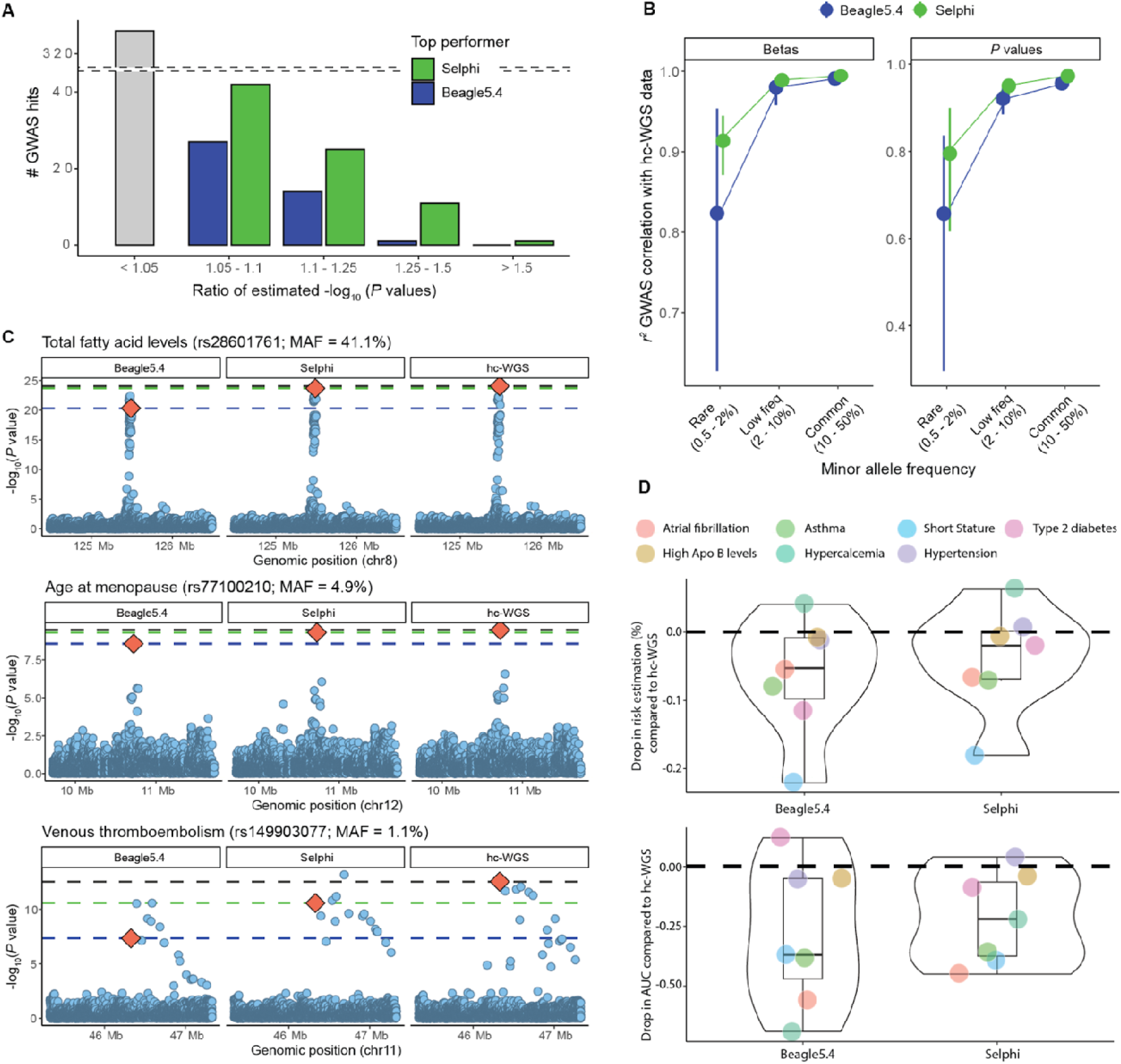
GWAS and PRS power analysis. (A) Number of GWAS hits in which Selphi or Beagle5.4 obtained higher significance, plotted by ratio bin. Variants that surpassed GWAS suggestive threshold (P < 10^-5^) were analyzed. A ratio below 1.05 was considered as an equivalent result for both Beagle5.4 and Selphi. (B) Squared correlation (*r*^2^) for betas and *P* values obtained from imputed sets and compared to hc-WGS across 50 UK biobank phenotypes by MAF. Nominally significant (P < 0.05) trait-associated hits collected by the GWAS Catalog were retrieved. Lower and upper limits of the forest plot represent the confidence interval from bootstrap resampling. (C) GWAS examples of imputed sets along with hc-WGS results for total fatty acid levels, age at menopause and venous thromboembolism phenotypes. Red diamond indicates known GWAS signals. (D) PRS drop in accuracy when comparing imputed sets with hc-WGS, assessed through relative risk and area under the curve (AUC).

## Discussion

Genotype imputation will likely continue to be an important part of future genomic studies, especially as large-population-wide genotyping efforts expand and reference panels continue to grow. Researchers will increasingly be able to impute and re-impute a larger number of rare variants and impute them to a higher quality. We have developed Selphi, a new software for genotype imputation that enables just that: improved overall and rare variant imputation. We benchmarked Selphi’s performance against three widely used imputation methods: Beagle5.4, Minimac4 and IMPUTE5 using the 1000 Genomes Project and TOPmed datasets. Selphi achieved better imputation accuracy than any other tested method, across all minor allele frequencies and ancestral backgrounds. In addition, we demonstrated Selphi’s utility in large biobank-scale datasets by showing Selphi’s superiority compared to the next most accurate model, Beagle5.4, using the UK Biobank dataset.

One of the main challenges in imputing rare variants is the lack of a suitable or large enough reference panel for accurate imputation (Sengupta et al. 2023, Terao et al. 2023) Selphi may help partially overcome this by implementing a heuristic IBD selection. By employing a rigorous selection protocol of haplotype selection, Selphi effectively prioritizes haplotypes that are more likely to share a true genetic lineage, as indicated by IBD, while reducing the likelihood of confounding IBS instances. This fine-tuned approach lays the groundwork for a more accurate and reliable imputation, which is particularly crucial when dealing with large genomic datasets where the precision of haplotype matching can significantly impact the overall imputation outcomes.

Unlike recent efforts to broaden and manipulate the reference panel (Mitt et al. 2017, Sun et al. 2022, Xu et al. 2022, Wuttke et al. 2023), Selphi increases accuracy within existing reference panels, which is of particular importance when there is not enough data to manipulate or broaden reference panels, which is often the case in non-European and non-British populations. Notably, Selphi achieved particularly pronounced improvements in East Asian and African populations of the 1000 Genomes Project and Chinese Americans in the Topmed dataset. These findings suggest that Selphi holds considerable promise for increasing imputation accuracy in populations that have been historically underrepresented in genetic research (Petrovski and Goldstein 2016, Martin et al. 2019, Atutornu et al. 2023).

It has been shown that improving imputation accuracy by improving the reference panel translates into improved downstream analysis (Huerta-Chagoya et al. 2023, Terao et al. 2023). Here we demonstrate that Selphi can improve both GWAS variant discovery and PRS calculation without changing the reference panel, obtaining results in closer alignment to hc-WGS.

In conclusion, Selphi is a promising genotype imputation method that achieves higher accuracy than existing methods, which can be used to boost downstream analyses, such as GWAS variant discovery and PRS. This advance in imputation, therefore, has the potential to improve the accuracy and resolution of future genomic studies.

## Acknowledgments

We thank the team and our colleagues at Omics Edge and Genius Labs. This research was conducted by using the UK Biobank Resource under application number 84038.

## Author contributions

Conceptualization and methodology, A.D.M. and P.G.Y.; data curation, A.D.M, A.A.M., S.B., A.T. and S.L.; formal analysis, A.D.M, A.A.M., S.B., J.L.J and S.L.; software, A.D.M, A.A.M., S.B., C.M. and M.M.; investigation, A.D.M, A.A.M., S.B., J.L.J, B.N. and C.M.; visualization, A.D.M., A.A.M., S.B., J.L.J and B.N.; writing - original draft, A.D.M, A.A.M. and B.N.; writing - review and editing, A.D M, S.B., J.L.J, B.N., S..L. and A.T.; funding acquisition, project administration, resources and supervision, P.G.Y and A.T.

## Data availability

The complete documented Selphi code is available in the following GitHub repository: https://github.com/selfdecode/rd-imputation-selphi. Additionally, we offer an applet that allows users to conveniently test the Selphi code on the UKB RAP platform.

All publicly available datasets utilized in this study can be accessed through their original publications and via application to the UK Biobank. Furthermore, access to The Trans-Omics for Precision Medicine (TOPMed) dataset can be granted by the National Institutes of Health (NIH).

