## Supplementary Material for "Selphi: Empowering GWAS Discovery through Enhanced Genotype Imputation"

Supplementary Data

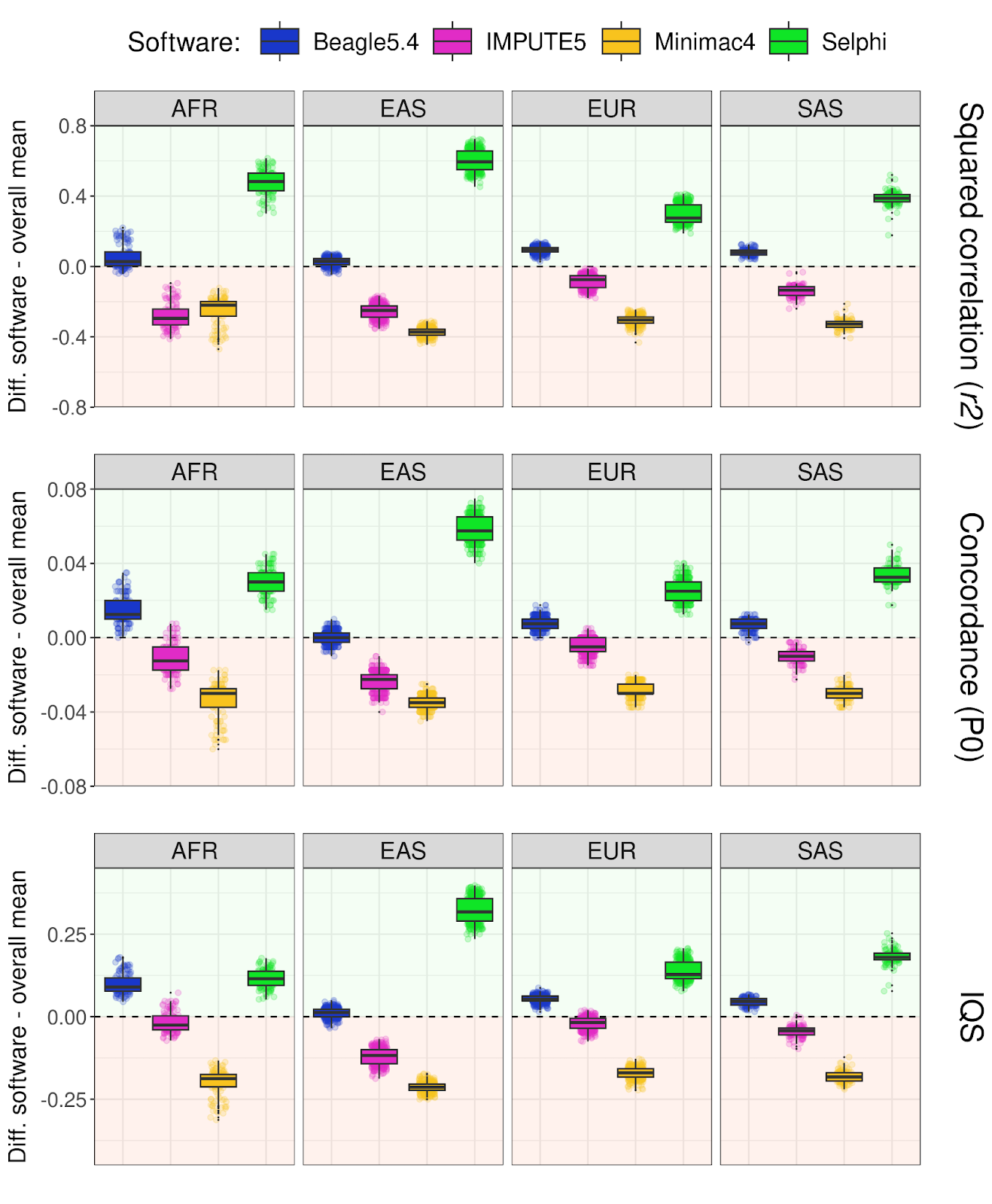

**Figure S1** **Imputation accuracy measured by different metrics in the 1000 Genomes Project dataset**. Difference in squared correlation, concordance, and F-score between Beagle5.4 (blue), IMPUTE5 (magenta), Minimac4 (yellow), and Selphi (green) for chromosomes 1-22 across different super-populations. The difference is shown as the deviation from the average number of errors across all four methods.

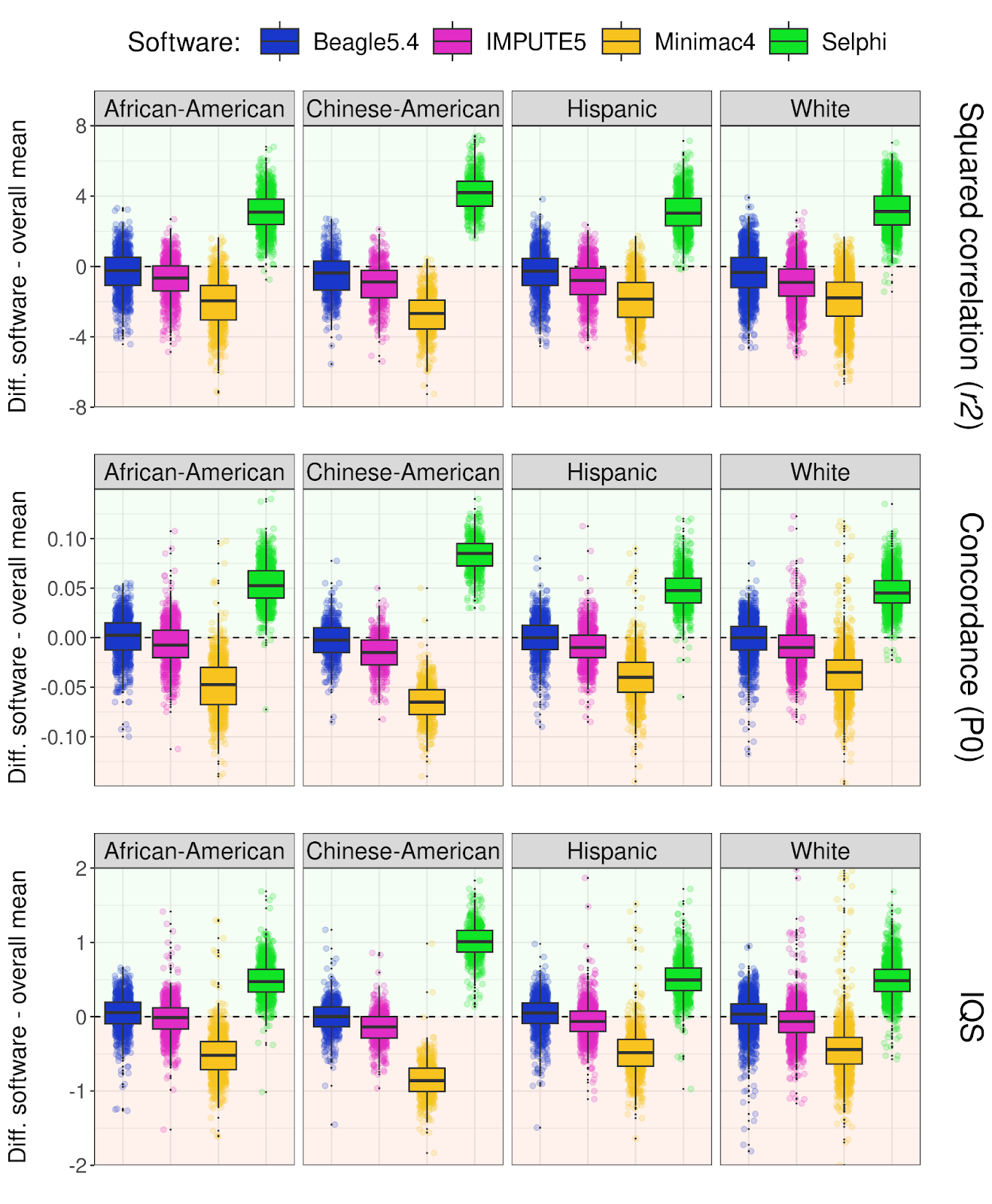

**Figure S2**  **Imputation accuracy measured by different metrics in the TOPMed dataset**. Difference in squared correlation, concordance, and F-score between Beagle5.4 (blue), IMPUTE5 (magenta), Minimac4 (yellow), and Selphi (green) for chromosome 20 across different super-populations. The difference is shown as the deviation from the average number of errors across all four methods.

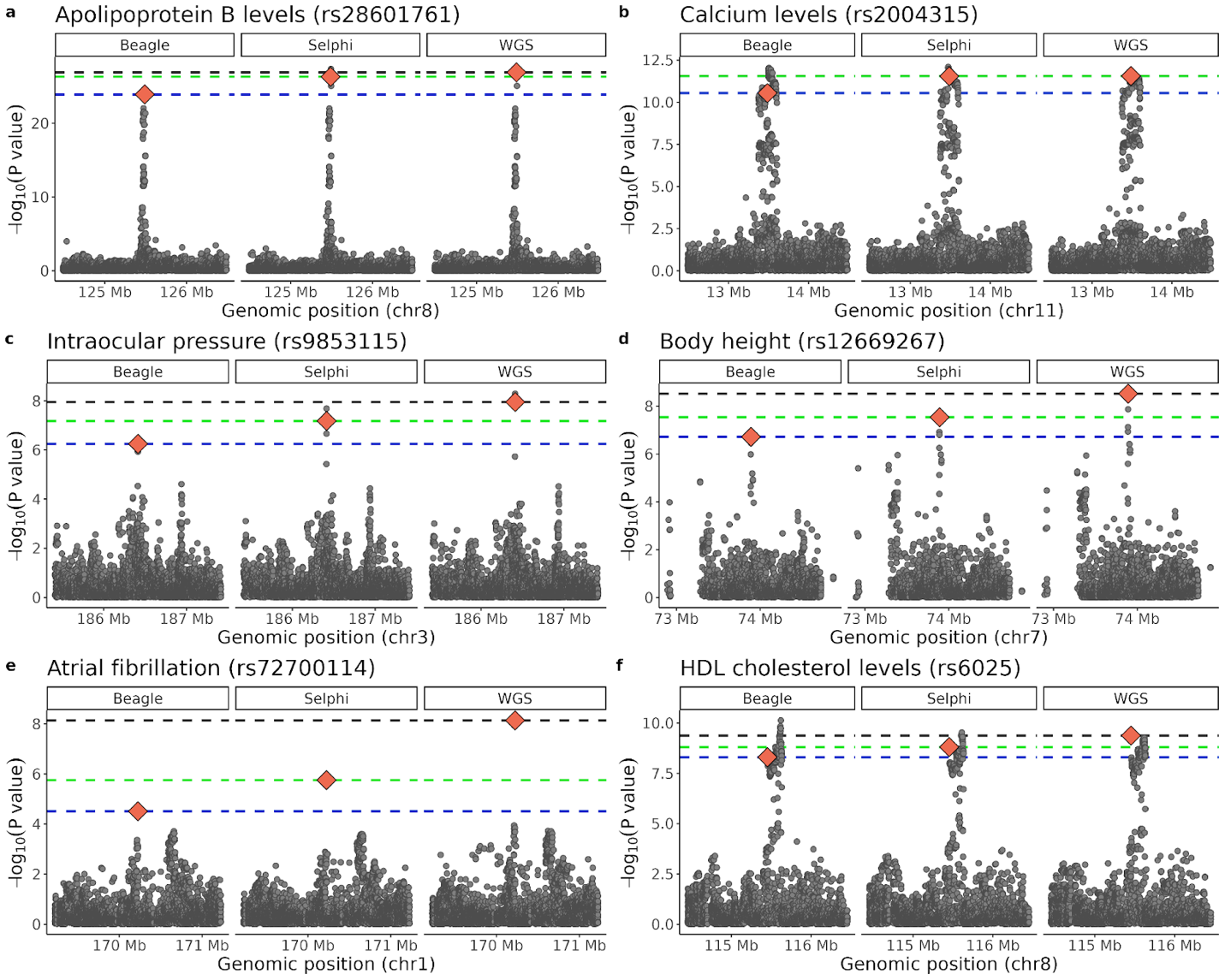
**Figure S3 Additional GWAS examples of imputed sets along with hc-WGS results.** Red diamond indicates the known GWAS signal collected by the GWAS Catalog and the horizontal lines, the significance achieved by Beagle5.4 (blue), Selphi (green) and hc-WGS (black) for these.

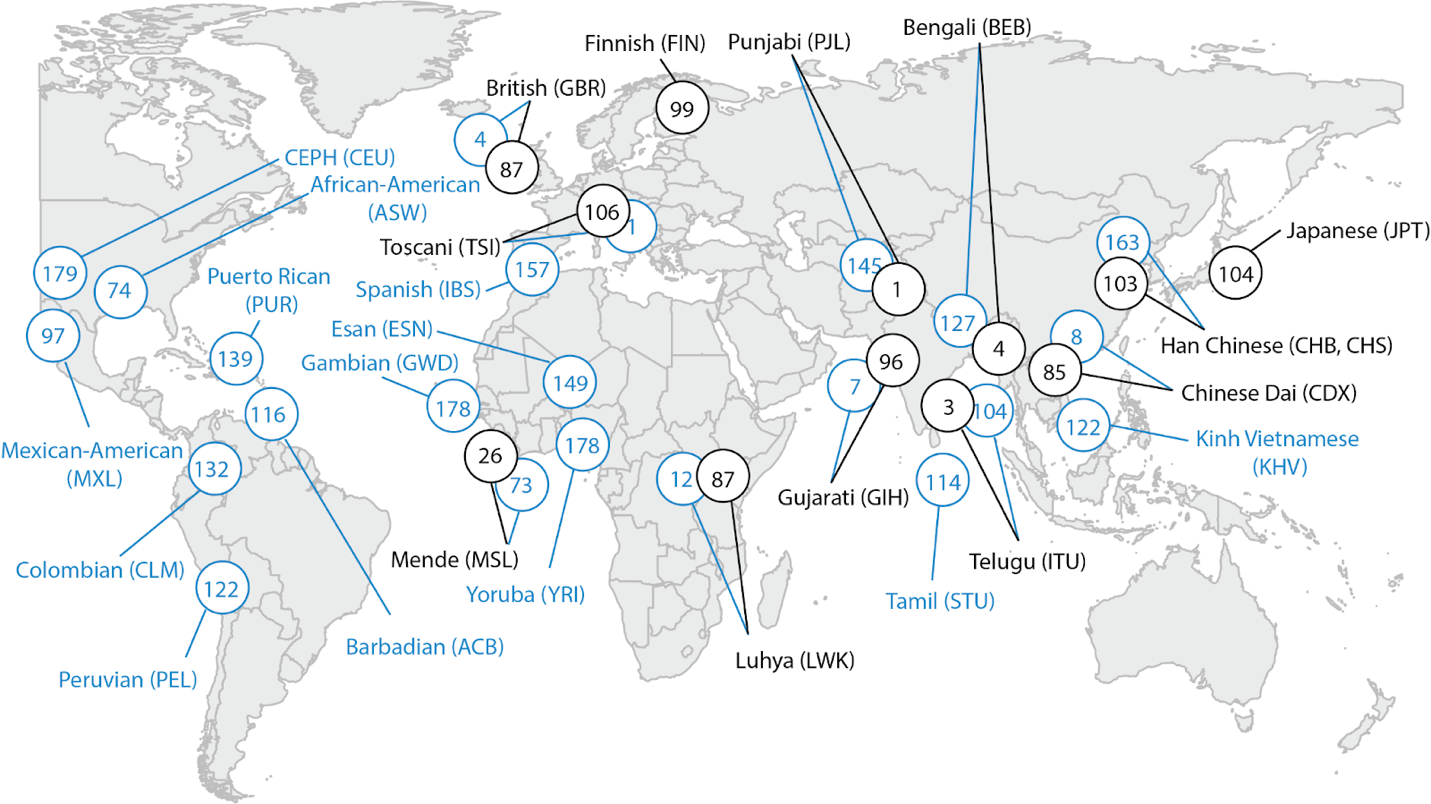

**Figure S4 Map of 1KGP samples.**

Samples used as imputation targets (black) and reference (blue) in the 1000 Genomes 30x Dataset.

**Table S1**. **Overall imputation accuracy in the 1KG, TOPMed and UK Biobank dataset for Beagle5.4, Minimac4, IMPUTE5 and Selphi.**

| **Method** | **Sum error** | **P_0_** | **IQS** | **R^2^** | **Dataset (N)** |
| --- | --- | --- | --- | --- | --- |
| **Selphi** | 383223835 | 0.99469 | 0.97042 | 0.95217 | 1KG (801) |
| **Beagle5.4** | 406946100 | 0.99436 | 0.96880 | 0.94827 |  |
| **IMPUTE5** | 420801415 | 0.99417 | 0.96776 | 0.94581 |  |
| **Minimac4** | 434165650 | 0.99399 | 0.96644 | 0.94438 |  |
| **Selphi** | 159396771 | 0.99449 | 0.91917 | 0.90559 | TOPMed (5000) |
| **Beagle5.4** | 175047669 | 0.99395 | 0.91388 | 0.86904 |  |
| **IMPUTE5** | 177332074 | 0.99387 | 0.91310 | 0.86430 |  |
| **Minimac4** | 187571372 | 0.99352 | 0.90843 | 0.85221 |  |
| **Selphi** | 9393206346 | 0.99984 | 0.98840 | 0.98079 | UKB (49997) |
| **Beagle5.4** | 11878020614 | 0.99980 | 0.98538 | 0.97673 |  |

**Table S2**. **Imputation accuracy for different MAF intervals in the 1KG dataset for Selphi, Beagle5.4, IMPUTE5, and Minimac4.**

| **MAF (%)** | **Method** | **Sum error** | **Mean error** | **Variants** | **P_0_** | **IQS** | **R^2^** |
| --- | --- | --- | --- | --- | --- | --- | --- |
| **0.05 - 0.1** | Selphi | 8960994 | 1.9251 | 4654733 | 0.9988 | 0.6191 | 0.3463 |
|  | Beagle5.4 | 9244895 | 1.9861 | 4654733 | 0.9988 | 0.6685 | 0.3632 |
|  | Minimac4 | 9052844 | 1.9449 | 4654733 | 0.9988 | 0.6552 | 0.3606 |
|  | IMPUTE5 | 9524045 | 2.0461 | 4654733 | 0.9987 | 0.6698 | 0.3573 |
| **0.1 - 0.2** | Selphi | 8255011 | 2.3240 | 3552051 | 0.9986 | 0.6729 | 0.4104 |
|  | Beagle5.4 | 8694171 | 2.4476 | 3552051 | 0.9985 | 0.7161 | 0.4215 |
|  | Minimac4 | 8469267 | 2.3843 | 3552051 | 0.9985 | 0.7025 | 0.4197 |
|  | IMPUTE5 | 9018244 | 2.5389 | 3552051 | 0.9984 | 0.7163 | 0.4149 |
| **0.2 - 0.5** | Selphi | 18865400 | 3.0244 | 6237778 | 0.9981 | 0.7448 | 0.4950 |
|  | Beagle5.4 | 20327351 | 3.2587 | 6237778 | 0.9980 | 0.7718 | 0.4967 |
|  | Minimac4 | 19826985 | 3.1785 | 6237778 | 0.9980 | 0.7598 | 0.4953 |
|  | IMPUTE5 | 21145176 | 3.3899 | 6237778 | 0.9979 | 0.7705 | 0.4892 |
| **0.5 - 1** | Selphi | 17934858 | 4.3390 | 4133453 | 0.9973 | 0.8134 | 0.5858 |
|  | Beagle5.4 | 19645450 | 4.7528 | 4133453 | 0.9970 | 0.8235 | 0.5781 |
|  | Minimac4 | 19254968 | 4.6583 | 4133453 | 0.9971 | 0.8150 | 0.5768 |
|  | IMPUTE5 | 20401500 | 4.9357 | 4133453 | 0.9969 | 0.8218 | 0.5703 |
| **1 - 2** | Selphi | 23401560 | 6.5277 | 3584963 | 0.9959 | 0.8534 | 0.6561 |
|  | Beagle5.4 | 25659883 | 7.1576 | 3584963 | 0.9955 | 0.8570 | 0.6438 |
|  | Minimac4 | 25375478 | 7.0783 | 3584963 | 0.9956 | 0.8497 | 0.6404 |
|  | IMPUTE5 | 26580136 | 7.4143 | 3584963 | 0.9954 | 0.8552 | 0.6357 |
| **2 - 5** | Selphi | 39724055 | 10.7033 | 3711382 | 0.9933 | 0.8931 | 0.7450 |
|  | Beagle5.4 | 43071817 | 11.6053 | 3711382 | 0.9928 | 0.8934 | 0.7307 |
|  | Minimac4 | 43954308 | 11.8431 | 3711382 | 0.9926 | 0.8854 | 0.7229 |
|  | IMPUTE5 | 44563655 | 12.0073 | 3711382 | 0.9925 | 0.8916 | 0.7229 |
| **5 - 10** | Selphi | 41439113 | 18.2768 | 2267308 | 0.9886 | 0.9283 | 0.8330 |
|  | Beagle5.4 | 44399138 | 19.5823 | 2267308 | 0.9878 | 0.9272 | 0.8213 |
|  | Minimac4 | 46775909 | 20.6306 | 2267308 | 0.9871 | 0.9201 | 0.8127 |
|  | IMPUTE5 | 45943670 | 20.2635 | 2267308 | 0.9874 | 0.9256 | 0.8149 |
| **10 - 20** | Selphi | 64286410 | 27.1053 | 2371727 | 0.9831 | 0.9498 | 0.8878 |
|  | Beagle5.4 | 68206516 | 28.7582 | 2371727 | 0.9821 | 0.9482 | 0.8801 |
|  | Minimac4 | 74156988 | 31.2671 | 2371727 | 0.9805 | 0.9422 | 0.8715 |
|  | IMPUTE5 | 70597872 | 29.7664 | 2371727 | 0.9814 | 0.9469 | 0.8753 |
| **20 - 50** | Selphi | 140734177 | 34.5483 | 4073546 | 0.9784 | 0.9622 | 0.9232 |
|  | Beagle5.4 | 148008323 | 36.3340 | 4073546 | 0.9773 | 0.9604 | 0.9183 |
|  | Minimac4 | 167406692 | 41.0961 | 4073546 | 0.9743 | 0.9553 | 0.9097 |
|  | IMPUTE5 | 153176542 | 37.6028 | 4073546 | 0.9765 | 0.9590 | 0.9147 |

**Table S3**. **Imputation accuracy for different MAF intervals in the TOPMed dataset for Selphi, Beagle5.4, IMPUTE5, and Minimac4.**

| **MAF (%)** | **Method** | **Sum error** | **Mean error** | **Variants** | **P_0_** | **IQS** | **R^2^** |
| --- | --- | --- | --- | --- | --- | --- | --- |
| **0.05 - 0.1** | Selphi | 812659 | 5.4131 | 150127 | 0.9995 | 0.7273 | 0.4362 |
|  | Beagle5.4 | 957775 | 6.3798 | 150127 | 0.9994 | 0.7432 | 0.4129 |
|  | IMPUTE5 | 1049218 | 6.9889 | 150127 | 0.9993 | 0.7433 | 0.4016 |
|  | Minimac4 | 984881 | 6.5603 | 150127 | 0.9993 | 0.7448 | 0.4102 |
| **0.1 - 0.2** | Selphi | 1121655 | 9.1522 | 122556 | 0.9991 | 0.7571 | 0.4835 |
|  | Beagle5.4 | 1344587 | 10.9712 | 122556 | 0.9989 | 0.7675 | 0.4564 |
|  | IMPUTE5 | 1473895 | 12.0263 | 122556 | 0.9988 | 0.7661 | 0.4423 |
|  | Minimac4 | 1379459 | 11.2557 | 122556 | 0.9989 | 0.7688 | 0.4531 |
| **0.2 - 0.5** | Selphi | 2393638 | 17.3295 | 138125 | 0.9983 | 0.7858 | 0.5404 |
|  | Beagle5.4 | 2906679 | 21.0438 | 138125 | 0.9979 | 0.7918 | 0.5121 |
|  | IMPUTE5 | 3224081 | 23.3418 | 138125 | 0.9977 | 0.7884 | 0.4941 |
|  | Minimac4 | 2991317 | 21.6566 | 138125 | 0.9978 | 0.7921 | 0.5077 |
| **0.5 - 1** | Selphi | 3077417 | 33.7669 | 91137 | 0.9966 | 0.8075 | 0.5915 |
|  | Beagle5.4 | 3772653 | 41.3954 | 91137 | 0.9959 | 0.8111 | 0.5620 |
|  | IMPUTE5 | 4231687 | 46.4322 | 91137 | 0.9954 | 0.8058 | 0.5400 |
|  | Minimac4 | 3884027 | 42.6175 | 91137 | 0.9957 | 0.8111 | 0.5573 |
| **1 - 2** | Selphi | 5050375 | 64.3278 | 78510 | 0.9936 | 0.8135 | 0.6158 |
|  | Beagle5.4 | 6226335 | 79.3063 | 78510 | 0.9921 | 0.8167 | 0.5861 |
|  | IMPUTE5 | 6965125 | 88.7164 | 78510 | 0.9911 | 0.8114 | 0.5644 |
|  | Minimac4 | 6397638 | 81.4882 | 78510 | 0.9919 | 0.8167 | 0.5820 |
| **2 - 5** | Selphi | 10559924 | 136.8362 | 77172 | 0.9863 | 0.8129 | 0.6226 |
|  | Beagle5.4 | 13021569 | 168.7344 | 77172 | 0.9831 | 0.8153 | 0.5945 |
|  | IMPUTE5 | 14540765 | 188.4202 | 77172 | 0.9812 | 0.8099 | 0.5745 |
|  | Minimac4 | 13395264 | 173.5767 | 77172 | 0.9826 | 0.8153 | 0.5911 |
| **5 - 10** | Selphi | 13591477 | 280.8272 | 48398 | 0.9719 | 0.8191 | 0.6441 |
|  | Beagle5.4 | 16480263 | 340.5154 | 48398 | 0.9659 | 0.8203 | 0.6142 |
|  | IMPUTE5 | 18217330 | 376.4067 | 48398 | 0.9624 | 0.8147 | 0.5955 |
|  | Minimac4 | 16908940 | 349.3727 | 48398 | 0.9651 | 0.8203 | 0.6109 |
| **10 - 20** | Selphi | 23206500 | 463.7405 | 50042 | 0.9536 | 0.8491 | 0.7065 |
|  | Beagle5.4 | 26833127 | 536.2121 | 50042 | 0.9464 | 0.8463 | 0.6765 |
|  | IMPUTE5 | 29339167 | 586.2909 | 50042 | 0.9414 | 0.8390 | 0.6566 |
|  | Minimac4 | 27375850 | 547.0575 | 50042 | 0.9453 | 0.8459 | 0.6733 |
| **20 - 50** | Selphi | 96561175 | 948.566 | 101797 | 0.9051 | 0.8223 | 0.6537 |
|  | Beagle5.4 | 100260273 | 984.904 | 101797 | 0.9015 | 0.8151 | 0.6337 |
|  | IMPUTE5 | 105134487 | 1032.7857 | 101797 | 0.8967 | 0.8067 | 0.6166 |
|  | Minimac4 | 100723193 | 989.4515 | 101797 | 0.9011 | 0.8139 | 0.6310 |

**Table S4**. **Imputation accuracy for different MAF intervals in the UK Biobank dataset for Beagle5.4 and Selphi.**

| **MAF (%)** | **Method** | **Sum error** | **Mean error** | **Total variants** | **P_0_** | **IQS** | **R^2^** |
| --- | --- | --- | --- | --- | --- | --- | --- |
| **0.05 - 0.1** | Selphi | 195 241 576 | 34.3962 | 5 676 254 | 0.9997 | 0.6893 | 0.3953 |
|  | Beagle5.4 | 224 017 164 | 39.4657 | 5 676 254 | 0.9996 | 0.7042 | 0.3376 |
| **0.1 - 0.2** | Selphi | 258 188 215 | 62.0330 | 4 162 113 | 0.9994 | 0.7435 | 0.4835 |
|  | Beagle5.4 | 289 180 235 | 69.4792 | 4 162 113 | 0.9993 | 0.7556 | 0.4159 |
| **0.2 - 0.5** | Selphi | 485 746 254 | 132.2627 | 3 672 586 | 0.9987 | 0.7930 | 0.5733 |
|  | Beagle5.4 | 536 055 465 | 145.9613 | 3 672 586 | 0.9985 | 0.8037 | 0.5068 |
| **0.5 - 1** | Selphi | 474 401 772 | 260.0186 | 1 824 492 | 0.9974 | 0.8528 | 0.6787 |
|  | Beagle5.4 | 535 200 645 | 293.3423 | 1 824 492 | 0.9971 | 0.8577 | 0.6243 |
| **1 - 2** | Selphi | 538 875 280 | 360.4527 | 1 494 996 | 0.9964 | 0.9102 | 0.7873 |
|  | Beagle5.4 | 631 960 858 | 422.7174 | 1 494 996 | 0.9958 | 0.9078 | 0.7475 |
| **2 - 5** | Selphi | 826 352 064 | 439.0366 | 1 882 194 | 0.9956 | 0.9539 | 0.8801 |
|  | Beagle5.4 | 1 018 628 170 | 541.1919 | 1 882 194 | 0.9946 | 0.9483 | 0.8541 |
| **5 - 10** | Selphi | 893 335 348 | 574.1385 | 1 555 958 | 0.9943 | 0.9730 | 0.9265 |
|  | Beagle5.4 | 1 129 591 446 | 725.9781 | 1 555 958 | 0.9927 | 0.9677 | 0.9095 |
| **10 - 20** | Selphi | 1 399 782 308 | 724.2896 | 1 932 628 | 0.9928 | 0.9809 | 0.9488 |
|  | Beagle5.4 | 1 814 799 973 | 939.0322 | 1 932 628 | 0.9906 | 0.9758 | 0.9363 |
| **20 - 50** | Selphi | 3 409 688 553 | 898.8476 | 3 793 400 | 0.9910 | 0.9844 | 0.9626 |
|  | Beagle5.4 | 4 549 889 687 | 1 199.4226 | 3 793 400 | 0.9880 | 0.9793 | 0.9532 |

**Table S5**. **TOPMed Studies and Consensus Groups** utilized for the construction of the TOPMed reference panel.

|  | **Study Name** | **Study ID** | **Consensus Group** | **Consensus Code** | **Version** |
| --- | --- | --- | --- | --- | --- |
| 1 | AfricanAm Sarcoidosis | phs001207 | c1 | DS-SAR-IRB | v2.p1 |
| 2 | ARIC WholeGene | phs001211 | c1 | HMB-IRB | v3.p2 |
| 3 | ARIC WholeGene | phs001211 | c2 | DS-CVD-IRB | v3.p2 |
| 4 | Asthma Barbados | phs001143 | c1 | GRU-IRB | v4.p1 |
| 5 | Asthma CostaRica | phs000988 | c1 | DS-ASTHMA-IRB-MDS-RD | v5.p1 |
| 6 | BrazilSCD | phs001468 | c1 | GRU-IRB-PUB-NPU | v1.p1 |
| 7 | CARDIA | phs001612 | c1 | HMB-IRB | v1.p1 |
| 8 | CARDIA | phs001612 | c2 | HMB-IRB-NPU | v1.p1 |
| 9 | CardioHealth | phs001368 | c1 | HMB-MDS | v2.p2 |
| 10 | CardioHealth | phs001368 | c2 | HMB-NPU-MDS | v2.p2 |
| 11 | CardioHealth | phs001368 | c4 | DS-CVD-NPU-MDS | v2.p2 |
| 12 | CardioHealth Amish | phs000956 | c2 | HMB-IRB-MDS | v4.p1 |
| 13 | Cleveland Family | phs000954 | c1 | DS-HLBS-IRB-NPU | v3.p2 |
| 14 | Cleveland Afib | phs001189 | c1 | GRU-IRB | v4.p1 |
| 15 | COPD | phs000951 | c1 | HMB-MDS | v4.p4 |
| 16 | CVHealth | phs000993 | c2 | DS-CVD-IRV-MDS | v5.p2 |
| 17 | CVHealth | phs000993 | c1 | HMB-IRB-MDS | v5.p2 |
| 18 | Framingham WholeGene | phs000974 | c1 | HMB-IRB-MDS | v4.p3 |
| 19 | Framingham WholeGene | phs000974 | c2 | HMB-IRB-NPU-MDS | v4.p3 |
| 20 | GeneSTAR | phs001218 | c2 | DS-CVD-IRB-NPU-MDS | v2.p1 |
| 21 | GENOA | phs001345 | c1 | DS-ASC-RF-NPU | v2.p1 |
| 22 | GenSalt | phs001217 | c1 | DS-HCR-IRB | v2.p1 |
| 23 | GOLDN | phs001359 | c1 | DS-CVD-IRB | v3.p1 |
| 24 | HCHS SOL | phs001395 | c1 | HMB-NPU | v2.p1 |
| 25 | HCHS SOL | phs001395 | c2 | HMB | v2.p1 |
| 26 | HyperGEN | phs001293 | c1 | GRU-IRB | v2.p1 |
| 27 | HyperGEN | phs001293 | c2 | DS-CVD-IRB-RD | v2.p1 |
| 28 | Jackson Heart | phs000964 | c1 | HMB-IRB-NPU | v4.p1 |
| 29 | Jackson Heart | phs000964 | c2 | DS-FDO-IRB-NPU | v4.p1 |
| 30 | Jackson Heart | phs000964 | c3 | HMB-IRB | v4.p1 |
| 31 | Jackson Heart | phs000964 | c4 | DS-FDO-IRB | v4.p1 |
| 32 | MESA | phs001416 | c1 | HMB | v2.p1 |
| 33 | MESA | phs001416 | c2 | HMB-NPU | v2.p1 |
| 34 | MGH Afib | phs001062 | c1 | HMB-IRB | v5.p2 |
| 35 | MGH Afib | phs001062 | c2 | DS-AF-IRB-RD | v5.p2 |
| 36 | MyLifeOurFuture Hemophilia | phs001515 | c1 | HMB-PUB | v2.p2 |
| 37 | Novel Risk Afib Women | phs001040 | c1 | HMB | v5.p1 |
| 38 | Partners HCBiobank | phs001024 | c1 | HMB | v4.p1 |
| 39 | PharmHU | phs001466 | c1 | HMB | v1.p1 |
| 40 | PharmHU | phs001466 | c2 | DS-SCD-RD | v1.p1 |
| 41 | PharmHU | phs001466 | c3 | DS-SCD | v1.p1 |
| 42 | SAFHS CVD | phs001215 | c1 | DS-DHD-IRB-PUB-MDS-RD | v3.p2 |
| 43 | SARP | phs001446 | c1 | GRU | v1.p1 |
| 44 | Vanderbilt AFAbalation | phs000997 | c1 | HMB-IRB | v4.p2 |
| 45 | Vanderbilt Afib | phs001032 | c1 | GRU-IRB | v6.p2 |
| 46 | Venous Throm | phs001402 | c1 | GRU | v3.p1 |
| 47 | WHI | phs001237 | c1 | HMB-IRB | v2.p1 |
| 48 | WHI | phs001237 | c2 | HMB-IRB-NPU | v2.p1 |

**Table S6. Reference and Target Markers across datasets.** Number of reference and target markers for each chromosome in our benchmarking panel obtained from the 1000 Genomes Project (1KG) Dataset and UK Biobank.

|  | **1000 Genomes Project** | | **TOPMed** | | **UK Biobank** | |
| --- | --- | --- | --- | --- | --- | --- |
| **Chr** | **Reference** | **Target** | **Reference** | **Target** | **Reference** | **Target** |
| 1 | 5,769,087 | 49,977 | / | / | 48,448,838 | 53,087 |
| 2 | 6,095,976 | 52,869 | / | / | 50,617,221 | 52,599 |
| 3 | 4,986,824 | 43,376 | / | / | 43,235,013 | 44,223 |
| 4 | 4,878,537 | 40,435 | / | / | 39,818,105 | 41,193 |
| 5 | 4,539,890 | 37,574 | / | / | 36,802,991 | 38,728 |
| 6 | 4,317,093 | 45,433 | / | / | 35,576,415 | 45,844 |
| 7 | 4,140,924 | 34,937 | / | / | 35,081,174 | 35,910 |
| 8 | 3,888,893 | 32,669 | / | / | 33,856,820 | 33,628 |
| 9 | 3,169,328 | 26,871 | / | / | 26,935,065 | 29,101 |
| 10 | 3,499,286 | 31,064 | / | / | 29,351,162 | 32,610 |
| 11 | 3,425,446 | 31,148 | / | / | 29,825,047 | 33,110 |
| 12 | 3,335,036 | 30,267 | / | / | 28,658,800 | 31,413 |
| 13 | 2,512,948 | 22,288 | / | / | 21,123,329 | 22,131 |
| 14 | 2,294,933 | 19,904 | / | / | 19,395,462 | 21,192 |
| 15 | 2,111,611 | 19,197 | / | / | 17,802,705 | 20,846 |
| 16 | 2,366,114 | 20,815 | / | / | 19,960,418 | 23,773 |
| 17 | 2,075,523 | 18,196 | / | / | 17,334,935 | 22,032 |
| 18 | 1,965,907 | 18,085 | / | / | 16,663,631 | 19,243 |
| 19 | 1,672,929 | 14,481 | / | / | 13,181,709 | 18,934 |
| 20 | 1,647,102 | 15,171 | 17,900,635 | 15,365 | 13,780,208 | 17,180 |
| 21 | 1,004,437 | 8,521 | / | / | 7,949,357 | 9,785 |
| 22 | 1,070,401 | 9,283 | / | / | 8,255,855 | 10,792 |

**Table S7. Phenotypic traits used in GWAS analysis.** QT = quantitative trait, Disease = binary trait.

| **Trait** | **Code** | **Type** |
| --- | --- | --- |
| **Age at menopause** | **AAM** | **QT** |
| **Apolipoprotein A1** | **APOEA** | **QT** |
| **Apolipoprotein B** | **APOEB** | **QT** |
| **Body mass index** | **BMI** | **QT** |
| **Calcium** | **CAL** | **QT** |
| **Docosahexaenoic acid** | **DOA** | **QT** |
| **Glycated hemoglobin** | **HBA1C** | **QT** |
| **High density lipoprotein cholesterol** | **HDL** | **QT** |
| **Height** | **HEIGHT** | **QT** |
| **Intraocular pressure** | **IOP** | **QT** |
| **Low density lipoprotein cholesterol** | **LDL** | **QT** |
| **Omega-6 fatty acids** | **OSFA** | **QT** |
| **Omega-3 fatty acids** | **OTFA** | **QT** |
| **Phosphatidylcholines** | **PDCL** | **QT** |
| **Polyunsaturated fatty acids** | **PFA** | **QT** |
| **Phosphoglycerides** | **PHG** | **QT** |
| **Resting heart rate** | **RHR** | **QT** |
| **Remnant cholesterol (Non-HDL and Non-LDL cholesterol)** | **RMNC** | **QT** |
| **Sphingomyelins** | **SGM** | **QT** |
| **Total cholesterol** | **TCH** | **QT** |
| **Total fatty acids** | **TFA** | **QT** |
| **Total triglycerides** | **TTG** | **QT** |
| **Alzheimer's disease** | **AD** | **Disease** |
| **Atrial fibrillation** | **AF** | **Disease** |
| **Age-related macular degeneration** | **AMD** | **Disease** |
| **Asthma** | **AST** | **Disease** |
| **Breast cancer** | **BC** | **Disease** |
| **Bipolar disorder** | **BD** | **Disease** |
| **Coronary artery disease** | **CAD** | **Disease** |
| **Crohn's disease** | **CD** | **Disease** |
| **Coeliac disease** | **CED** | **Disease** |
| **Bowel cancer** | **CRC** | **Disease** |
| **Cardiovascular disease** | **CVD** | **Disease** |
| **Epithelial ovarian cancer** | **EOC** | **Disease** |
| **Hypertension** | **HT** | **Disease** |
| **Ischaemic stroke** | **ISS** | **Disease** |
| **Melanoma** | **MEL** | **Disease** |
| **Multiple sclerosis** | **MS** | **Disease** |
| **Osteoporosis** | **OP** | **Disease** |
| **Prostate cancer** | **PC** | **Disease** |
| **Parkinson's disease** | **PD** | **Disease** |
| **Primary open angle glaucoma** | **POAG** | **Disease** |
| **Psoriasis** | **PSO** | **Disease** |
| **Rheumatoid arthritis** | **RA** | **Disease** |
| **Schizophrenia** | **SCZ** | **Disease** |
| **Systemic lupus erythematosus** | **SLE** | **Disease** |
| **Type 1 diabetes** | **T1D** | **Disease** |
| **Type 2 diabetes** | **T2D** | **Disease** |
| **Ulcerative colitis** | **UC** | **Disease** |
| **Venous thromboembolic disease** | **VTE** | **Disease** |

**Table S8.** Studies used for GWAS meta-analysis for each phenotype in PRS analysis.

| **Trait** | **Studies** |
| --- | --- |
| Height | [Yengo et al. 2022](https://www.nature.com/articles/s41586-022-05275-y) |
| Calcium | [Dennis et al. 2021](https://pubmed.ncbi.nlm.nih.gov/33441150/), [Sakaue et al. 2021](https://www.nature.com/articles/s41588-021-00931-x) |
| Atrial fibrillation | [Christophersen et al. 2017](https://www.nature.com/articles/ng.3843), [Sakaue et al. 2021](https://www.nature.com/articles/s41588-021-00931-x), [Kurki et al. 2023](https://doi.org/10.1038/s41586-022-05473-8) |
| Asthma | [Demenais et al. 2018](https://pubmed.ncbi.nlm.nih.gov/29273806/), [Sakaue et al. 2021](https://www.nature.com/articles/s41588-021-00931-x), [Kurki et al. 2023](https://doi.org/10.1038/s41586-022-05473-8) |
| Apolipoprotein B | [Kettunen et al. 2016](https://www.nature.com/articles/ncomms11122), [Sun et al. 2018](https://www.nature.com/articles/s41586-018-0175-2), [Gudjonsson et al. 2022](https://www.nature.com/articles/s41467-021-27850-z), [Thareja et al. 2023](https://pubmed.ncbi.nlm.nih.gov/36168886/) |
| Hypertension | [Wojcik et al. 2019](https://pubmed.ncbi.nlm.nih.gov/31217584/), [Kurki et al. 2023](https://doi.org/10.1038/s41586-022-05473-8) |
| Type 2 diabetes | [Wojcik et al. 2019](https://pubmed.ncbi.nlm.nih.gov/31217584/), [Vujkovic et al. 2020](https://pubmed.ncbi.nlm.nih.gov/32541925/), [Kurki et al. 2023](https://doi.org/10.1038/s41586-022-05473-8) |
